# Functional trajectory following pediatric stroke: a cohort study of acute inpatient rehabilitation outcomes

**DOI:** 10.1101/2024.01.11.24301187

**Authors:** Jennifer Wu, Deena S. Godfrey, Patricia Orme, Brian D. Wishart

## Abstract

**Background:** Stroke in childhood is a significant cause of morbidity and mortality. Neurologic impairments due to childhood stroke are associated with long-term disability and decreased quality of life. However, there are limited studies examining functional outcomes of childhood stroke. The goal of this study was to characterize functional outcomes of children and adolescents admitted to acute inpatient rehabilitation following stroke.

**Methods:** A retrospective cross-sectional study of 100 patients aged 0 to 21 years admitted to a pediatric acute inpatient rehabilitation program following new diagnosis of stroke in childhood. The primary outcome measures were Functional Independence Measure in Children (WeeFIM) score at admission and discharge. Secondary outcome measures included change in WeeFIM score and IRF Efficiency score.

**Results:** The 56 male/43female/1 transgender patients were 10.4±6.1 years old with ischemic (n=53), hemorrhagic (n=41), and hemorrhagic converted ischemic (n=6) strokes. At admission, the group demonstrated moderate-to-severe functional impairments (WeeFIM total score=47.9±26.3 points). Inpatient rehabilitation length of stay was 34.1±28.6 days and at inpatient rehabilitation discharge, WeeFIM total score improved to 73.2±31.5 points, representing a group IRF Efficiency score of 1.42±1.5 points/day. Group effects were also found for medical management of agitation, stroke prophylaxis, and stimulant therapy.

**Conclusions:** Acute inpatient rehabilitation demonstrates statistically and clinically significant functional improvements following pediatric stroke as measured on the WeeFIM scale. Additional studies are needed to examine group effects found from medical management in the inpatient rehabilitation setting.

## INTRODUCTION

Stroke in childhood is a significant cause of mortality and morbidity^1^. In North America, annual incidence of childhood stroke ranges from 1.2 to 13 cases per 100,000 and some studies report mortality rates in childhood stroke as high as 54%^2,3^. Among survivors of childhood stroke, chronic neurologic impairments occur in nearly 50% of childhood stroke survivors and have long-term effects on physical, psychosocial, and academic outcomes^4,5^. While stroke is a major cause of long-term disability acquired in childhood, there are limited studies examining functional outcomes of childhood stroke^6–8^.

Regarding stroke type and functional outcomes, pediatric stroke is distinct compared to stroke in adults. Among children and adolescents, hemorrhagic stroke occurs at significantly higher rates compared to stroke in adults and overall outcomes following pediatric stroke are generally more favorable with a greater proportion of children and adolescents achieving normative function compared to adult stroke survivors^9–12^. These differences highlight etiologic variability in pediatric and adult stroke and therefore limit attempts to extrapolate results from studies on stroke outcomes in adult populations to pediatric populations^13^. Thus, specific studies on functional outcomes after stroke in children and adolescents are needed.

Early rehabilitation trajectory following childhood stroke demonstrates some predictive effects on long-term outcomes^4,14^. Studies reporting functional outcomes in the acute inpatient rehabilitation setting in childhood stroke are limited but generally find inpatient rehabilitation improves stroke-related neurologic impairments in children and adolescents^14,15^. Additional studies that also examine functional outcomes with respect to stroke type, acute hospital course, and inpatient rehabilitation course may provide additional insights into variability across patients in response to inpatient rehabilitation and subsequent long-term functional outcomes.

To broaden the limited pediatric stroke rehabilitation literature, this study examines functional outcomes in children and adolescents admitted to acute inpatient rehabilitation for neurologic impairments following new diagnosis of stroke. Additional analyses included group effects with respect to stroke type, acute hospital course, and inpatient rehabilitation course on functional outcomes. This study is also the first to report on the effects of stroke prophylaxis and stimulant therapy on functional outcomes in the inpatient rehabilitation setting.

## METHODS

### Study Design and Population

This was a retrospective cross-sectional study of patients aged 0 to 21 years who were admitted to a pediatric unit at a free-standing rehabilitation hospital between January 1, 2017 and July 31, 2023. Potential participants were identified through chart review. Stroke diagnosis was confirmed through review of clinical and radiographic exams. All patients with new, radiologically confirmed diagnosis of stroke during the preceding acute hospital admission were included in the study. All study procedures were approved by the Institutional Review Board of Mass General Brigham. Due to the retrospective study design, this study was exempted from written and verbal consent procedures. The STROBE checklist (www.strobe-statement.org) reporting guideline was followed.

### Procedures

Patient data including demographics, disease etiology, functional impairments, radiographic and laboratory studies, and acute care and rehabilitation hospital course were obtained from the medical records. The Functional Independent Measure in Children (WeeFIM) was used to track functional improvements across the inpatient rehabilitation admission^16^. The WeeFIM is a 126-point ordinal scale which assesses function across 18 items rated on a scale of 1 (dependent) to 7 (independent) the domains of self-care, mobility, and cognition (**Table 1**). WeeFIM data were extracted from facility-specific listings in the Uniform Data System for Medical Rehabilitation (UDSMR). The primary outcomes measures were WeeFIM score at admission and discharge. To account for age-related effects on WeeFIM scoring, age-corrected WeeFIM scores were calculated from a product of the actual WeeFIM score and a ratio of the age-expected WeeFIM score divided by the overall maximum WeeFIM score^17^. Secondary outcome measures were change in uncorrected and age-corrected WeeFIM scores from admission to discharge and IRF(Inpatient Rehabilitation Facility) Efficiency score. The IRF Efficiency score was calculated from the ratio of change in WeeFIM score from admission to discharge and acute inpatient rehabilitation length of stay and expressed as points/day^18^.

**Table 1:**
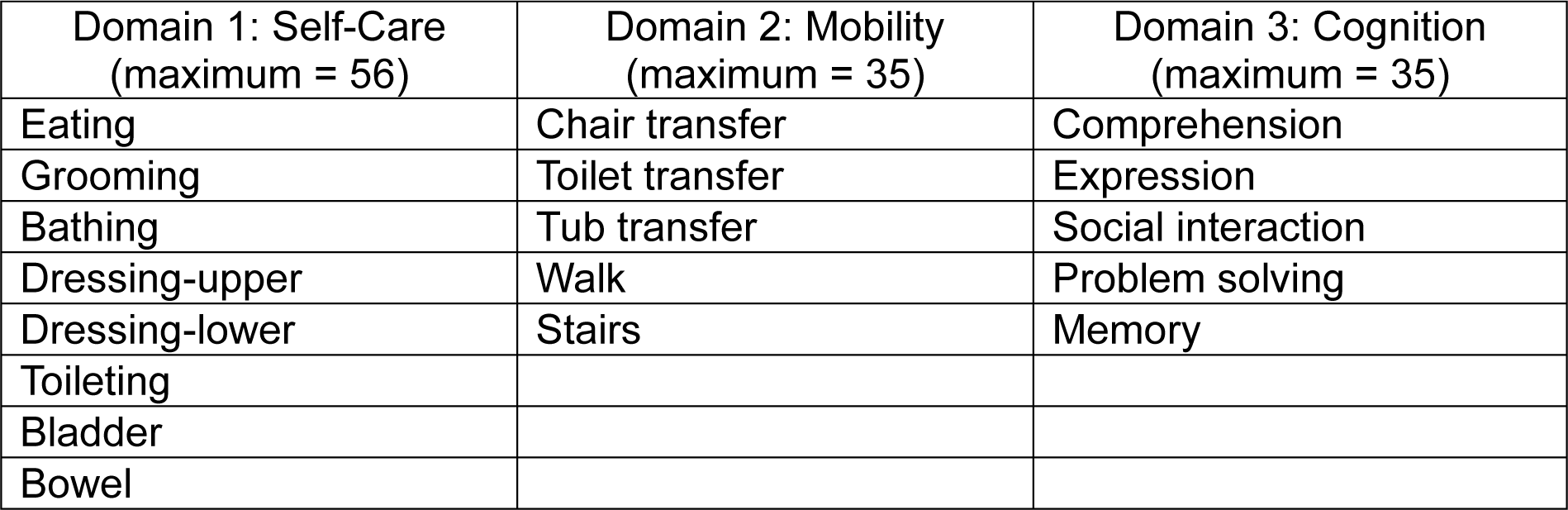
Functions scored for each domain of the WeeFIM scale.

### Statistics

Shapiro-Wilks testing found uncorrected and age-corrected WeeFIM total and domain-specific scores were non-normal. Therefore, nonparametric statistical methods were used throughout. Continuous variables were evaluated with Spearman rank order test and categorical variables were evaluated with Wilcoxon signed rank order test. Contingency testing to compare categorical variables was performed using Fisher’s exact test. All analyses were 2-tailed with alpha=0.05, Bonferroni corrected, and performed with Matlab R2015a (MathWorks Inc, Natick, MA).

## RESULTS

### Patient cohort

The group of 100 infants, children, and adolescents were 56 male/43 female/1 transgender with mean age=10.4±6.1 years. They ranged 0.5 to 19.8 years old (median [IQR]=11.0 [4.8-16.2] years) at admission to acute inpatient rehabilitation. There were no perinatal strokes in the group. There were 12 patients who had a previous inpatient rehabilitation admission at an outside facility prior to transferring care. The group also included 3 patients with 2 separate inpatient rehabilitation admissions for different stroke injuries.

Rupture of arteriovenous malformation was the most common etiology for inpatient rehabilitation admission for stroke (n=32). Moya-Moya disease and Moya-Moya like syndrome (n=11), malignancy (n=10), congenital heart disease (n=8), vasculitis (n=7), and dissection (n=6) were also frequent etiologies of stroke. There were 20 patients who had stroke in the setting of medical-surgical complications during their acute hospital course, which included infection, seizure, hypoxia due to respiratory disease, hypoperfusion, and hypercoagulability. Ten patients had cryptogenic stroke and otherwise negative workup.

There were more ischemic (n=53) compared to hemorrhagic strokes (n=41) in this group with a smaller group of ischemic converted to hemorrhagic stroke (n=6). The patients with ischemic stroke was younger (9.5±6.1 years) compared to the group with hemorrhagic stroke (11.5±5.8 years, **Figure 1**). A small group were admitted with repeat stroke (n=7). Most patients had unilateral stroke affecting the Left brain (n=44); 34 patients had unilateral stroke affecting the Right brain; and 22 patients had bilateral strokes. While stroke injuries spanned all cerebral and cerebellar vascular distributions, stroke affecting the middle cerebral artery distribution was the most common (n=48).

**Figure 1.**
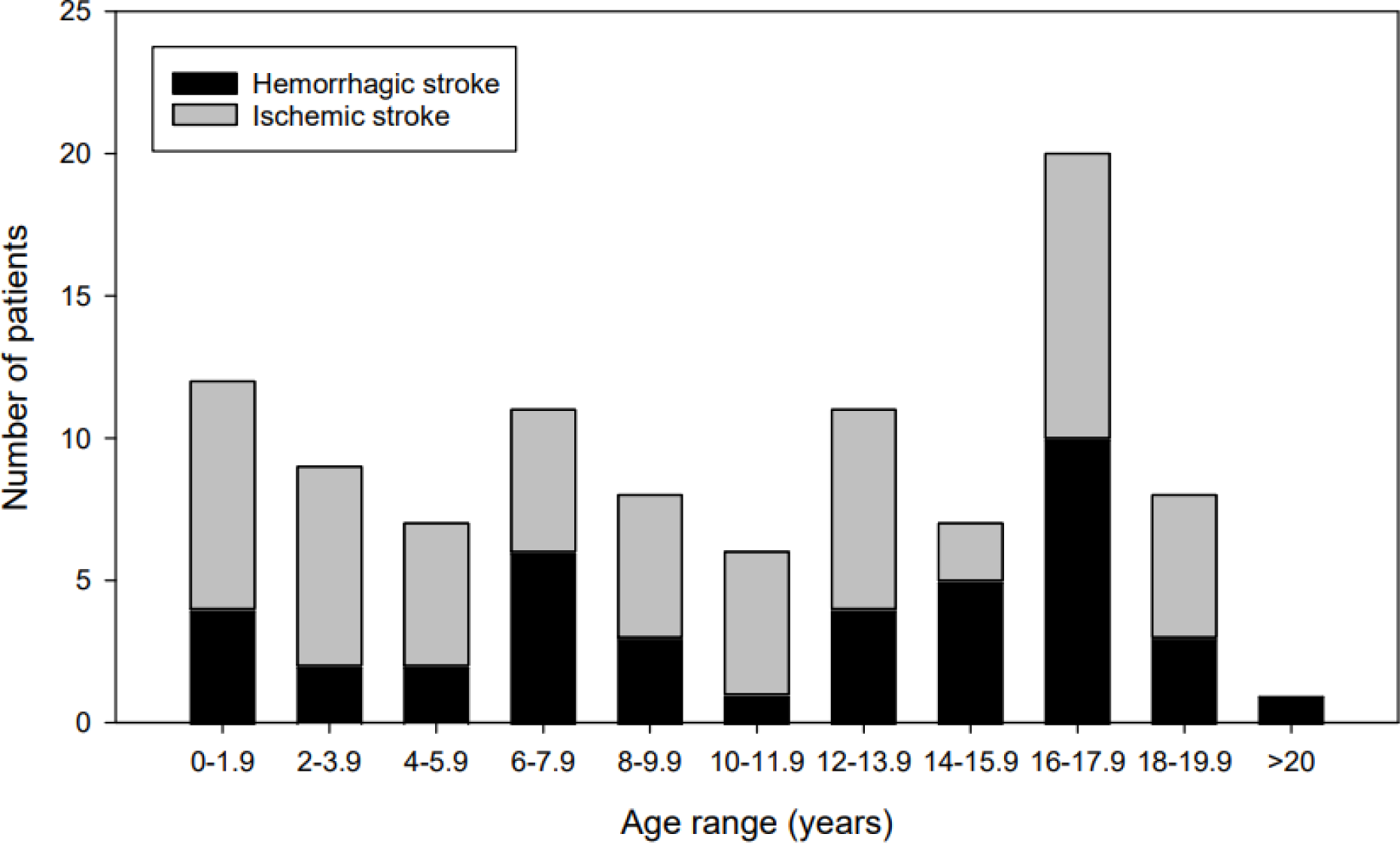
Histogram of pediatric stroke patient ages at acute inpatient rehabilitation admission by stroke type.

### Acute hospital course

The acute hospital length of stay preceding inpatient rehabilitation admission was 33.2±35.7 (median[IQR]=23[12-40]) days. Neurosurgical interventions included decompressive craniectomy in 23 patients, ventriculoperitoneal (VP) shunt placement in 13 patients, and lobectomy in 5 patients. There were 6 patients who had both decompressive craniectomy and VP shunt placement. Two patients had decompressive craniectomy, VP shunt placement, and lobectomy. Acute hospital length of stay for patients who had decompressive craniectomy (56.7±56.6 days) was longer compared to patients who did not (26.1±22.6 days, Z=3.29, *p*=0.001). Patients who had VP shunt placement (71.8±70.9 days) also had longer acute hospital length of stay compared to patients who did not (27.4±22.4 days, Z=2.85, *p*=0.004). The average length of stay for patients who had lobectomy (69.0±105.5 days) was longer than patients who did not (31.3±28.2 days), but this difference did not achieve significance.

### Inpatient rehabilitation course

At admission to inpatient rehabilitation, the group demonstrated functional impairments across all domains. The admission WeeFIM total score was 47.9±26.3 points (maximum:126 points) with domain-specific scores 21.2±12.8 points in Self-Care (maximum: 56 points), 11.1±6.3 points in Mobility (maximum: 25 points), and 15.6±9.7 points in Cognition (maximum: 25 points). Among patients aged ≥ 2 years at admission (n=88), nine patients were dependent with wheelchair and the remaining 79 patients were walking at least short-distances with variable requirements for assistance. With respect to feeding, 58 patients were exclusively taking nutrition by mouth, 19 patients had a nasogastric tube, and 23 patients had a gastrostomy tube in place; there was 1 patient who had a gastrostomy tube at baseline prior to new stroke diagnosis. Among the 76 patients who had been cleared for an oral diet order, 42 patients were admitted with a dysphagia diet order.

The inpatient rehabilitation length of stay was 34.1±28.6 days which ranged from 3 to 134 days (median[IQR]=23.5[14-42] days). There were 80 patients who had a single, continuous inpatient rehabilitation course without leave-of-absence. For the 20 patients who had a leave of absence, average leave of absence was 13.3±21.9 days. The group achieved statistically significant improvement in both uncorrected and age-corrected WeeFIM scores from admission to discharge (**Figure 2**). At discharge, uncorrected total WeeFIM score was 73.2±31.5 points with average improvement 25.3±17.4 points. The discharge uncorrected WeeFIM domains scores were 32.6±15.7 points for Self-Care, 19.6±8.3 points for Mobility, and 20.9±9.5 points for Cognition with an improvement of 11.4±9.1 points, 8.6±6.0 points, and 5.3±5.0 points, respectively. There were no patients who experienced a decline in uncorrected total WeeFIM score across the inpatient rehabilitation course; five patients did not improve uncorrected total WeeFIM score at discharge. Among patients ≥3 years at admission (n=79), age-corrected WeeFIM total score was 55.3±25.0 points at admission and improved to 86.5±21.7 points at discharge with an improvement of 31.2±15.9 points. The group IRF Efficiency score was 1.42±1.5 (median[IQR]=1.24[0.7-1.6) points/day.

**Figure 2.**
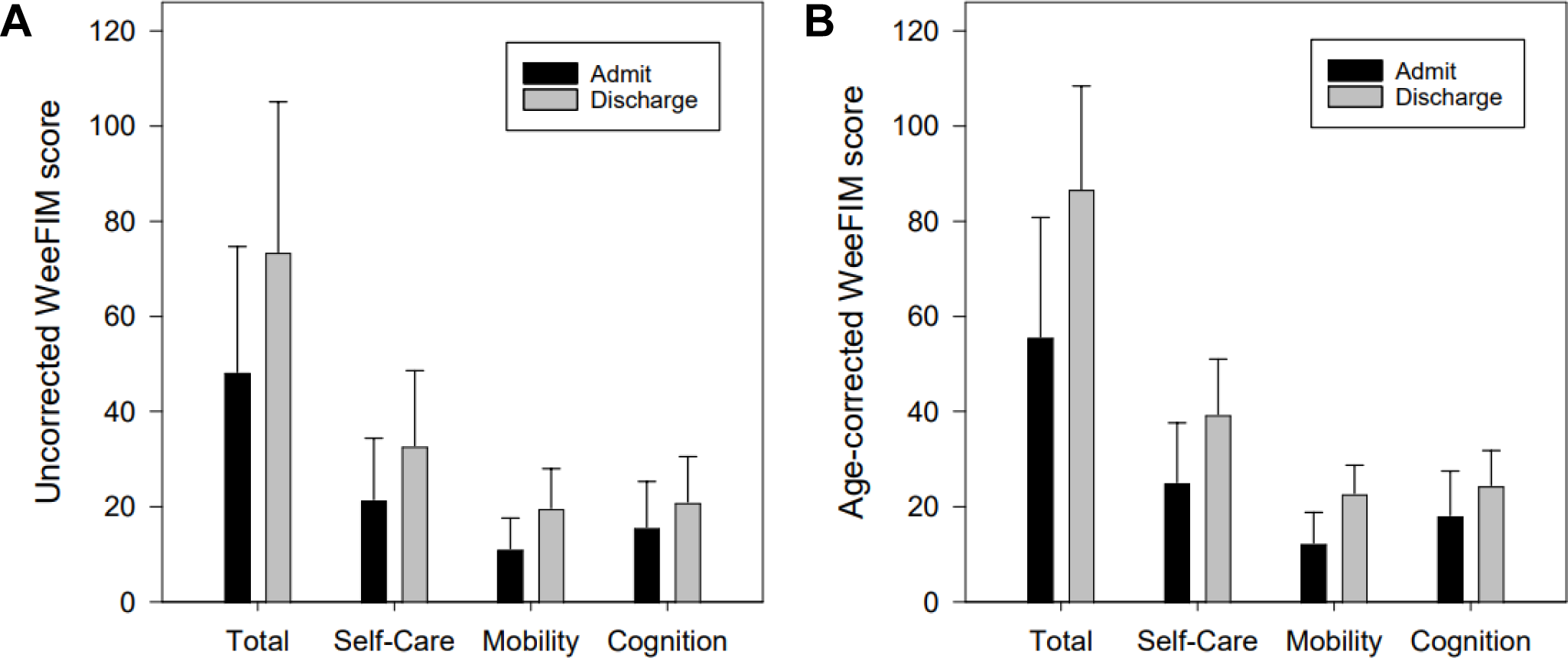
WeeFIM total and domain-specific scores at acute inpatient rehabilitation from admission to discharge. Group mean and standard deviation are demonstrated for uncorrected (A) and age-corrected (B) scores.

At discharge, most patients were ambulatory and taking an oral diet. There were 87 patients walking at discharge which included 8 patients who had improvement in mobility category from wheelchair-dependent to walking with assistive device and/or bracing. Feeding status also improved across the group at discharge. Seventy-two patients were discharged home on an oral diet, including 9 who had been admitted with a nasogastric tube in place and 6 who had a new gastrostomy tube placed during the preceding acute hospitalization. Of the patients who were admitted with a nasogastric tube in place, 6 were discharged home with ongoing requirement for the nasogastric tube and 4 were discharged home with a new gastrostomy tube placed during the inpatient rehabilitation admission. Among patients admitted with a gastrostomy tube in place, 52% of patients had improvement in their IDDSI (International Dysphagia Diet Standardisation Initiative) diet level, and among patients admitted with nasogastric tube in place, 63% had improvement in IDDSI diet level during inpatient rehabilitation.

### Effects of stroke type on hospital course and functional outcome

Patients with hemorrhagic stroke had a longer acute hospital length of stay compared to patients with ischemic stroke (hemorrhagic: 35.5±30.4 days; ischemic: 31.6±39.2 days; *Z*=2.25, *p*=0.03). Those with hemorrhagic stroke were also more likely to have history of craniectomy compared to patients with ischemic stroke (*p*=0.01). However, there was not a significant effect of stroke type on history of VP shunt placement or lobectomy during the acute hospital course. At admission, patients with hemorrhagic stroke had lower age-corrected WeeFIM score (49.0±24.5 points) compared to patients with ischemic stroke (60.4±24.6 points; *Z*=2.1, *p*=0.04), but this difference was not seen in age-corrected WeeFIM total scores at discharge. While stroke type did not have an effect on IRF Efficiency, patients with hemorrhagic stroke tended to have slightly longer inpatient rehabilitation admission length of stay (38.3±28.5 vs 31.1±28.5 days, *p*=0.08) and larger gains in age-corrected WeeFIM total score during the inpatient rehabilitation admission (34.5±16.7 vs 28.5±14.8 points, *p*=0.07).

### Age effects on stroke type, hospital course, and functional outcomes

There was not a significant effect of age on stroke type, acute hospital length of stay, or inpatient rehabilitation length of stay. There also was not a significant effect of age on history of craniectomy or lobectomy during the acute hospital course. However, patients who had VP shunt placement were younger than patients who did not have VP shunt placement (7.2±6.3 years vs 10.8±5.9 years; *Z*=2.1, *p*=0.04). Across the group, older age was correlated with higher uncorrected WeeFIM total score at admission (n=100, *r*=0.58, *p*<0.0001) and at discharge (*r*=0.78, *p*<0.0001). For patients aged ≥3 years, there was a weaker relationship between older age and higher age-corrected WeeFIM total score at admission (n=79, *r*=0.32, *p*=0.004) and at discharge (*r*=0.46, *p*<0.001). While older age showed a positive relationship with larger improvement in uncorrected WeeFIM total score across the inpatient rehabilitation course (*r*=0.54, *p*<0.0001), there was not a statistical relationship between age and improvement in age-corrected WeeFIM total score (*p*>0.3) or IRF Efficiency (*p*>0.8).

Age showed the largest effects on age-corrected WeeFIM scores in the Cognition domain and the smallest effects in the Mobility domain. At admission, older age was correlated with higher age-corrected WeeFIM scores in Self-Care (n=79, *r*=0.31, p=0.006) and Cognition (*r*=0.34, p=0.002) but not in Mobility (*p*>0.1). Similarly at discharge, older age was correlated with higher age-corrected WeeFIM scores in Self-Care (*r*=0.46, *p*<0.0001), Mobility (*r*=0.28, *p*=0.01), and Cognition domains (*r*=0.40, *p*=0.0003). There was not an effect of age on improvement in age-corrected WeeFIM domain scores in Self-Care (p>0.1), Mobility (p>0.3), or Cognition (p>0.7).

### Medical management in the inpatient rehabilitation setting

There were 34 patients requiring medication to treat agitation at inpatient rehabilitation admission. These patients were generally younger (8.2±5.8 vs 11.5±5.9 years, *p*=0.01) and had longer acute hospital length of stay (56.6±49.3 vs 20.9±16.0 days, *p*<0.0001). Compared to patients who were not on an agent to treat agitation, these patients also had lower age-corrected WeeFIM total score at inpatient rehabilitation admission (38.0±20.7 vs 62.1±23.4 points, *p*=0.0001). Those who had a history of craniectomy during the acute hospital course were more likely to be admitted to inpatient rehabilitation on an agent to treat agitation (*p*=0.005), but there was no difference in frequency of VP shunt placement or lobectomy during the acute hospital course. Patients who were discharged from inpatient rehabilitation with an agent to treat agitation had longer inpatient rehabilitation length of stay (45.7±23.6 vs 32.3±29.0 days, *p*=0.008), lower discharge age-corrected WeeFIM total score (60.1±25.8 vs 89.1±19.6 points, *p*=0.006), and lower IRF Efficiency (0.48±0.3 vs 1.52±1.6 points/day, *p*=0.0006).

At admission, 46 of 100 patients were on anticoagulant or antiplatelet therapy for secondary stroke prophylaxis. Most patients were on aspirin monotherapy (n=30) with aspirin/clopidogrel being the most common dual therapy (n=5). There were also patients on clopidogrel (n=2), warfarin (n=2), rivaroxaban (n=1), and apixaban (n=1) monotherapy; there were 2 patients on aspirin/enoxaparin dual therapy. Patients with ischemic stroke were more likely to be admitted to inpatient rehabilitation with anticoagulant or antiplatelet therapy compared to those with hemorrhagic stroke (ischemic: 43/59 vs hemorrhagic: 3/41, *p*<0.001). Across the group, patients admitted on anticoagulant or antiplatelet therapy had higher age-corrected admission WeeFIM total score (65.8±20.8 vs 46.8±25.1 points, *p*=0.001), shorter inpatient rehabilitation length of stay (42.4±34.4 vs 25.3±17.1 days, *p*=0.02), and higher IRF Efficiency (1.46±0.8 vs 1.40±1.9 points/day, *p*=0.04).

The most common medication class introduced during the inpatient rehabilitation admission were stimulants. In total, there were 24 patients who were initiated on and discharged home with new stimulant therapy for stroke-related attention impairments. The most common stimulant therapy at discharge was immediate-release methylphenidate monotherapy (n=17) followed by combination therapy with long-acting and immediate-release methylphenidate (n=4), long-acting methylphenidate monotherapy (n=2), and guanfacine monotherapy (n=1). The youngest patient who was trialed on stimulant therapy was 4.1 years-old, but there was not a significant effect of age on trial of stimulant therapy during inpatient rehabilitation. The acute hospital course was similar between patients who were and were not trialed on stimulant therapy during the inpatient rehabilitation course with respect to acute hospital length of stay and neurosurgical interventions (including craniectomy, VP shunt placement, and lobectomy). Patients who were trialed on stimulant therapy during the inpatient rehabilitation admission had lower age-corrected total WeeFIM score at admission (46.5±24.2 vs 61.2±24.1 points, *p*=0.01) with the largest differences noted in the Cognition domain (14.7±7.5 vs 20.5±9.8 points, *p*=0.01). At discharge from inpatient rehabilitation, patients who were trialed on stimulant therapy had similar age-corrected WeeFIM total score and inpatient rehabilitation length of stay compared to patients who had not been trialed on stimulant therapy. As a result, patients who were trialed on stimulant therapy trended towards higher IRF Efficiency (1.77±1.2 vs 1.3±1.7 points/day, *p*=0.07) compared to patients who were not trialed on stimulant therapy.

## DISCUSSION

The study found children and adolescents with new diagnosis of stroke demonstrated global and domain-specific functional improvements across a 34-day acute inpatient rehabilitation admission. As in previous reports, this group of children and adolescents with new diagnosis of stroke showed a slight predominance of males and demonstrated higher frequency of hemorrhagic stroke compared to adult populations^14,19^. Among this group demonstrating moderate-to-severe impairments following stroke, patient age, stroke type, and acute hospital course did not have significant effects on the inpatient rehabilitation course. While previous studies have capitalized on regional and national pediatric stroke registries to identify epidemiologic risk factors^9,19,20^, this study represents the largest single-site assessment of inpatient rehabilitation outcomes in pediatric stroke.

Children and adolescents admitted to inpatient rehabilitation with new stroke-related functional impairments demonstrated statistical and clinical gains as measured in repeated WeeFIM assessments. The group showed improvement in total WeeFIM score from 47.9 points at admission to 73.2 points at discharge, which correspond to clinical improvement from maximal assistance level of function at admission to moderate assistance level of function at discharge^17^. This group of pediatric stroke patients admitted to inpatient rehabilitation experienced WeeFIM improvements comparable to a mixed group of children with central neurologic impairments that included stroke, traumatic brain injury, and cancer ^21^. Compared to pediatric inpatient rehabilitation patients with nontraumatic hypoxic or anoxic brain injury, this group of pediatric stroke patients were less impaired at admission and demonstrated greater improvements during inpatient rehabilitation^22^. In contrast, a group of patients with traumatic brain injury had higher IRF Efficiency compared to this group of pediatric stroke patients^23^. These differences likely reflect higher medical complexity and extensive medical workup during the acute hospital course for pediatric stroke patients resulting in overall delayed admission to inpatient rehabilitation and greater global functional impairments at admission. Regardless, this group of pediatric stroke patients demonstrated statistically significant improvements in both global function, as reflect by change in WeeFIM total score, and domain-specific improvements in Self-Care, Mobility, and Cognition WeeFIM subscores. In addition, domain-specific improvements of 11.4 points in Self-Care, 8.6 points in Mobility, and 5.3 points in Cognition across the inpatient rehabilitation admission exceed previously published Minimum Clinically Important Differences in each WeeFIM domains-specific score^24^. Together, these results support acute inpatient rehabilitation to be a beneficial intervention for optimizing recovery trajectory in pediatric patients with new neurologic impairments following stroke.

In the inpatient rehabilitation setting, older age at admission was correlated with higher WeeFIM total score at both admission and discharge, but there was not a statistical relationship between age and change in WeeFIM total score. These results align with previous reports demonstrating older age is generally associated with better outcomes following pediatric stroke^9,25^. While there are several studies examining the effects of age on cognitive outcomes following pediatric stroke^25–27^, this study also reports age effects on stroke outcomes in both cognitive and non-cognitive functional domains in the inpatient rehabilitation setting. Similar to a previous smaller study evaluating functional outcomes of inpatient rehabilitation in children with stroke in Saudi Arabia^15^, this study also found age did not affect response to therapeutic intervention (or change in WeeFIM score from inpatient rehabilitation admission to discharge) following pediatric stroke. These findings suggest that all patients, regardless of age at stroke onset, can benefit from inpatient rehabilitation following pediatric stroke.

Pharmacologic treatment of agitation was associated with worse outcomes compared to patients who were not using medication to treat agitation. In this study of pediatric stroke, 34 of 100 patients were admitted with an agitation medication. In contrast, a multicenter study of adult stroke admissions to inpatient rehabilitation reported less than 10% of individuals required pharmacologic treatment for agitation^28^. This difference may be partially attributed to higher rates of hemorrhagic stroke in pediatric compared to adult populations as agitation is a more frequent symptom in hemorrhagic compared to ischemic stroke^29^. The frequency of an agent to treat agitation remains significantly higher in this pediatric stroke group than rates cited in the adult population in both hemorrhagic and ischemic stroke^28,30^. As in adult stroke, use of an agent to manage agitation in the inpatient rehabilitation setting was associated with longer lengths of stay and lesser WeeFIM gains in this group of pediatric stroke patients. Interestingly, the rates of medical management for agitation in this group of pediatric stroke patients are consistent with reported rates of agitation in traumatic brain injury^31^. Given strong associations between increased agitation and poor clinical outcomes in the inpatient rehabilitation setting in patients with traumatic brain injury^32^, the relatively high rates of agitation found in this study highlight the need for vigilant assessment and management of agitation following pediatric stroke.

Stroke prophylaxis including anticoagulation and antiplatelet therapy at inpatient rehabilitation admission was correlated with higher WeeFIM score at admission and higher IRF Efficiency. In adult stroke, aspirin and antiplatelet therapies have shown benefits with respect to decreasing mortality and recurrent stroke^33,34^. Although evidence for efficacy is sparse in pediatric populations, expert clinical opinions on clinical management extrapolated from adult stroke studies generally support recommendations to use aspirin and other antiplatelet agents for secondary stroke prophylaxis in children and adolescents^35,36^. To the best of our knowledge, this study is the first to examine the functional effects of stroke prophylaxis on inpatient rehabilitation outcomes in a pediatric population. Given that patients admitted on medical therapy for stroke prophylaxis demonstrated a lesser degree of impairment at inpatient rehabilitation admission and higher IRF Efficiency, the current study findings suggest antiplatelet and anticoagulant therapy may support an enhanced brain substrate for pediatric post-stroke rehabilitation. Pre-clinical studies have shown antiplatelet therapy improve cerebral blood flow in the neonatal and ischemia-affected brain which may be contributing to improved outcomes among patients receiving antiplatelet and anticoagulant therapy for stroke prophylaxis^33,37^. Additional studies are needed to define the mechanisms that underlie differences in functional outcomes found in this study with respect to secondary stroke prophylaxis.

Stimulant therapy may accelerate functional recovery among pediatric patients admitted to acute inpatient rehabilitation following stroke. Specifically, IRF Efficiency among patients who had stimulant trial during inpatient rehabilitation was 1.77 points/day compared to 1.3 points/day among patients who did not have stimulant trial. While the current study found only a trend of association, prospective follow-up studies that demonstrate differences in recovery trajectories with stimulant therapy in the inpatient rehabilitation setting could have significant long-term implications for academic and psychosocial outcomes following pediatric stroke^38,39^. With rates of attention disorders as high as 50 percent in some studies of pediatric stroke^39,40^, studies examining the short- and long-term effects of stimulant therapy for cognitive outcomes are especially salient for the pediatric stroke population. To date, literature supporting the use of stimulant therapy in the inpatient rehabilitation setting is primarily focused on pediatric traumatic brain injury^41,42^. The current results provide additional support demonstrating the use of medications such as amantadine and methylphenidate to treat disorders of attention and executive function following acquired brain injuries in childhood.

The current study is limited by its retrospective, cross-sectional design. Prospective, multi-center studies that confirm the findings are imperative for continuing to characterize the natural history, short- and long-term outcomes, and therapeutic targets for improving rehabilitation in pediatric stroke. Compared to the existing literature^14,15^, this study reports on inpatient rehabilitation outcomes for the largest single-site group of pediatric stroke patients to date. These findings provide baseline metrics for future studies examining the effects of novel therapeutic interventions. In addition, the group differences noted across patients who had differing medical management in the inpatient rehabilitation setting (with respect to stroke prophylaxis and stimulant therapy) represents potential new avenues for investigation both for improving clinical outcomes and for defining the physiologic mechanisms which underlie pediatric stroke rehabilitation. Prospective studies will be needed to determine the indication, optimal dosing, and duration of interventions to maximize functional trajectories following stroke in children and adolescents.

Pediatric stroke is an important cause of childhood-acquired disability. There are a wide range and variety of deficits that can result from pediatric stroke and these deficits have the potential to have lifelong implications across all aspects of health and wellness. As a result, recovery and outcomes are at the forefront of patients and parents’ minds from the first moments of injury and prognosis conversations at this stage can often be challenging. Reassuringly, the current study found acute inpatient rehabilitation to be a functionally beneficial intervention for pediatric patients with new neurologic impairments due to stroke and represent a benchmark from which clinicians can begin discussions on prognosis and expected outcomes. Interestingly, medical intervention in the inpatient rehabilitation setting demonstrated effects on length of stay and functional level at discharge from inpatient rehabilitation. Further investigations into the proactive medical treatment of stroke-related neurologic impairments, including agitation, attention impairment, and secondary stroke prophylaxis may further improve functional gains achieved in the inpatient rehabilitation setting.

## Data Availability

Subject deidentified data will be made available upon request at the discretion of the authors and in accordance with Mass General Brigham data sharing policies.

## ACKNOWLEDGMENTS

The authors thank DN for her critical review and editorial contributions to this manuscript.

